# Protective behaviours and secondary harms from non-pharmaceutical interventions during the COVID-19 epidemic in South Africa: a multisite prospective longitudinal study

**DOI:** 10.1101/2020.11.12.20230136

**Authors:** Guy Harling, Francesc Xavier Gómez-Olivé, Joseph Tlouyamma, Tinofa Mutevedzi, Chodziwadziwa Whiteson Kabudula, Ruth Mahlako, Urisha Singh, Daniel Ohene-Kwofie, Rose Buckland, Pedzisai Ndagurwa, Dickman Gareta, Resign Gunda, Thobeka Mngomezulu, Siyabonga Nxumalo, Emily B. Wong, Kathleen Kahn, Mark J. Siedner, Eric Maimela, Stephen Tollman, Mark Collinson, Kobus Herbst

**Affiliations:** Africa Health Research Institute, KwaZulu-Natal, South Africa; Institute for Global Health, University College London, London, United Kingdom; Department of Epidemiology & Harvard Center for Population and Development Studies, Harvard T.H. Chan School of Public Health, Boston, MA, USA; MRC/Wits Rural Public Health and Health Transitions Research Unit (Agincourt), School of Public Health, Faculty of Health Sciences, University of the Witwatersrand, Johannesburg, South Africa; DIMAMO Population Health Research Centre, School of Health Care Sciences, Faculty of Health Sciences, University of Limpopo, Limpopo, South Africa; Faculty of Science and Agriculture, School of Mathematical and Computer Sciences, Department of Computer Science, University of Limpopo, Limpopo, South Africa; DSI-MRC South African Population Research Infrastructure Network, South Africa; School of Nursing and Public Health, University of KwaZulu-Natal, Durban, South Africa; Division of Infectious Diseases, University of Alabama Birmingham, Birmingham, AL, USA; INDEPTH Network, Accra, Ghana; Harvard Medical School and the Medical Practice Evaluation Center, Massachusetts General Hospital, Boston, MA, USA; Department of Public Health, School of Health Care Sciences, Faculty of Health Sciences, University of Limpopo, Limpopo, South Africa

**Keywords:** lockdown, knowledge, behaviour change, economic wellbeing, mental health, healthcare access, rural, peri-urban

## Abstract

**Background:** In March 2020 South Africa implemented strict non-pharmaceutical interventions (NPIs) to contain Covid-19. Over the subsequent five months NPIs were eased in stages according to national strategy. Covid-19 spread throughout the country heterogeneously, reaching rural areas by July and peaking in July-August. Data on the impact of NPI policies on social and economic wellbeing and access to healthcare is limited. We therefore analysed how rural residents of three South African provinces changed their behaviour during the first epidemic wave.

**Methods:** The South African Population Research Infrastructure Network (SAPRIN) nodes in Mpumalanga (Agincourt), KwaZulu-Natal (AHRI) and Limpopo (DIMAMO) provinces conducted longitudinal telephone surveys among randomly sampled households from rural and peri-urban surveillance populations every 2-3 weeks. Interviews included questions on: Covid-19 knowledge and behaviours; health and economic impact of NPIs; and mental health.

**Results:** 2262 households completed 10,966 interviews between April and August 2020. By August, self-reported satisfaction with Covid-19 knowledge had risen from 48% to 85% and facemask use to over 95%. As selected NPIs were eased mobility increased, and economic losses and anxiety and depression symptoms fell. When Covid-19 cases spiked at one node in July, movement dropped rapidly, and missed daily medication rates doubled. Economic concerns and mental health symptoms were lower in households receiving a greater number of government-funded old-age pensions.

**Conclusions:** South Africans reported complying with stringent Covid-19 NPIs despite the threat of substantial social, economic and health repercussions. Government-supported social welfare programmes appeared to buffer interruptions in income and healthcare access during local outbreaks. Epidemic control policies must be balanced against impacts on wellbeing in resource-limited settings and designed with parallel support systems where they threaten income and basic service access.

## BACKGROUND

Shortly after the global epidemic of novel coronavirus disease 2019 (Covid-19) was declared a Public Health Emergency of International Concern, South Africa was identified as highly vulnerable due to: i) extensive transport and airlinks [1]; ii) burden of infectious and non-communicable health conditions [2, 3]; and iii) large socioeconomically vulnerable population [4]. National government rapidly announced strict nationwide non-pharmaceutical interventions (NPIs) (“Level 5 lockdown”) on 26^th^ March, 2020. Under these NPIs leaving home was only allowed for groceries, medicines/medical care or to conduct permitted essential work, tobacco and alcohol sales were banned and from 1^st^ May facemask use was mandatory in public spaces. These regulations were accompanied with guidance on enhanced use of hand washing or sanitizer and surface cleaning.

The lockdown was intended to: (i) reduce Covid-19 transmission through strictly restricting physical interaction; (ii) avoid rapid epidemic growth and allow healthcare providers to prepare for a subsequent rise in demand for care; (iii) promote widespread educational campaigns to reduce Covid-19 transmission; and (iv) initiate an ambitious, country-wide, community-based Covid-19 screening and testing programme [5]. Between May and September the lockdown was gradually eased, allowing return to work, school, and limited public gatherings.

While cumulative Covid-19 cases grew slowly during April, the incidence curve accelerated from May onwards, peaking between June and August. By 1^st^ September 2020, South Africa had reported over 625,000 confirmed cases and over 14,000 deaths [6], among the 10 largest reported epidemics worldwide at that date [7]. The true impact of epidemic appears to be even greater, with 42,396 excess deaths reported between January and August nationally, compared to 2018-19 [8].

Relaxation of lockdown regulations, even as the epidemic curved upwards, reflected competing health and economic vulnerabilities and priorities, alongside sustained popular pressure. Concern was widespread that the lockdown was substantially affecting the national economy and individual household livelihoods, as well as access to education, healthcare and medication [4, 9]. Additionally, some expected lockdown to be futile since much of the population could not maintain physical distancing or implement NPIs, due to household and community overcrowding and limited access to running water and sanitation [10].

Robust data are essential to evaluate these competing hypotheses, and to target resources where they are most needed. Although South Africa has effective national healthcare surveillance systems, there is limited capacity to monitor social and behavioural effects of NPIs on the Covid-19 epidemic at a local level. NPIs such as those implemented in South Africa might be expected to bring differing risks and benefits across socioeconomic settings. For example, in rural areas unease surrounded water access for hand hygiene and “imilindo” (funeral night vigils held in crowded rooms) [11], while urban areas worried about dwelling proximity and shared ablutions [12]. To date, most data collected on these impacts have been limited to online or urban settings, with only one including longitudinal follow-up [13-16]. Robust comparison between urban and rural settings is limited yet vital if public sector responses are to be effectively aligned with prevailing conditions.

We leveraged an existing research infrastructure in three South African provinces to evaluate how health, social and economic behaviours changed as regulations and the national epidemic changed from April to August 2020.

## METHODS

### Study site

The South African Population Research Infrastructure Network (SAPRIN) is an initiative hosted by the South African Medical Research Council and funded long-term by the National Department of Science and Innovation. SAPRIN integrates three Health and Demographic Surveillance System (HDSS) nodes for population and health surveillance: (i) the MRC/Wits Rural Public Health and Health Transitions Research Unit (Agincourt) in Ehlanzeni district, Mpumalanga [17]; (ii) DIMAMO Population Health Research Centre in Capricorn district, Limpopo [18]; and (iii) the Africa Health Research Institute (AHRI) in uMkhanyakude district, KwaZulu-Natal [19]. Further urban nodes are under development. The nodes, each containing over 100,000 individuals residing in approximately 20,000 households, vary in settlement structure and density. The three districts are rural or peri-urban, located in the east of South Africa (Supplementary Figure 1) and poor relative to the country. Nodes conduct multiple in-person and telephonic surveys per year to update health and sociodemographic data, although DIMAMO had only partially captured socioeconomic data for the first time before Covid-19 arrived.

### Study design

In March 2020, SAPRIN initiated plans for each HDSS node to implement a high-intensity, longitudinal telephonic survey covering at least 750 randomly selected households in each setting, using telephone numbers extracted from each node’s most recent census. This sample was selected to provide estimates of survey- and wave-specific proportions with precision of no less than 4 percentage points, assuming 80% response rate in line with past SAPRIN surveys. Every 2-3 weeks, a primary respondent was called from a central call centre at each node and invited to answer questions on behalf of the household. Individual-level questions could be answered by other household members if they were present; otherwise, the primary respondent served as a proxy. The questionnaire included Covid-19 symptom screening. Individuals meeting Department of Health symptom criteria for Covid-19 were referred for further investigation, possible testing and care. Telephone calls continued from April to August with continuous quality monitoring. Implementation at AHRI has been described in detail elsewhere [20].

### Measures

This analysis focuses on questions relating to responses to Covid-19 and NPIs in three key domains: 1) Covid-19 knowledge and behaviour; 2) health and economic impacts of NPIs; and 3) mental health. For behaviour, the primary respondent was asked about their perceived knowledge about Covid-19 on a five-point scale; we separated respondents into those self-reporting less than enough knowledge vs. those reporting enough or more. Respondents were then asked about household behaviour change in response to the Covid-19 epidemic. Questions included whether any resident household member had left the house in the past seven days, and whether any non-household members visited the house during the preceding day; we divided respondents into those reporting any vs. those reporting none for each question. Respondents were also asked if household members had avoided crowded areas or social events, travel (using local minibus taxis or long-distance travel) and if they used facemasks when going out.

For health and economic impact, primary respondents were asked about household members’ ability to: (i) access all needed daily medications; (ii) access needed healthcare; and (iii) earn money. Finally, for mental health we asked primary respondents the Generalized Anxiety Disorder 2-item (GAD-2) and Patient Health Questionnaire 2-item (PHQ-2) questions. We scored both GAD-2 and PHQ-2 as positive for scores of 3 or higher using standard, South African validated, cut-offs [21].

We linked data from this high-intensity survey to routine individual and household socio-demographic data collected in 2019 on household: number of children, working-age adults and pension-age adults; maximal educational attainment; node-specific asset index quintile; level of employment and unconditional social grant receipt.

### Statistical analysis

We included in this analysis anyone interviewed before 1^st^ September 2020. We first described survey response rates across each node and time period, as well as key pre-Covid-19 household characteristics. We next described change by node and month of data collection for each of eleven measures across the three domains (behaviour; health and economic impact; mental health).

We then ran multivariable regression models to assess independent predictors of our outcomes of interest, using complete case analysis. For each outcome we fitted a Poisson model with household-level random effects and robust standard errors (generating prevalence ratios). Initially we included node, interview round and month of interview as covariates, and then added household characteristics. Data analysis was conducted in Stata v15.1 (StataCorp, College Station, TX, USA) and R v4.0.2 [22].

### Ethical considerations

All households had previously given consent to be contacted by phone and each primary respondent gave recorded, verbal consent. All study procedures were discussed with each nodes’ existing community advisory groups and approved by Limpopo, Mpumalanga and KwaZulu-Natal’s provincial Department of Health Research Ethics Committees (REC), the University of KwaZulu-Natal’s Biomedical REC, the University of the Witwatersrand’s Human REC (Medical) and the University of Limpopo’s Turfloop REC.

## RESULTS

Between 15^th^ April and 31^st^ August 2020, AHRI completed six waves of data collection, MRC/Wits-Agincourt five and DIMAMO two (Figure 1). These waves covered almost the entire period of the first Covid-19 epidemic wave in South Africa, including outbreaks of varying sizes in the three provinces under observation. Response rates averaged 78.7% with variation from 65.6% to 90.3% by wave and node (Figure 1). Direct refusal was rare (3.1% of all attempts), while unanswered calls were more common (10.7%) and numbers were quite often out of service or claimed to be wrong numbers (7.6%). In total, 10,966 household interviews were completed with 2,662 unique households between April and August.

**Figure 1:**
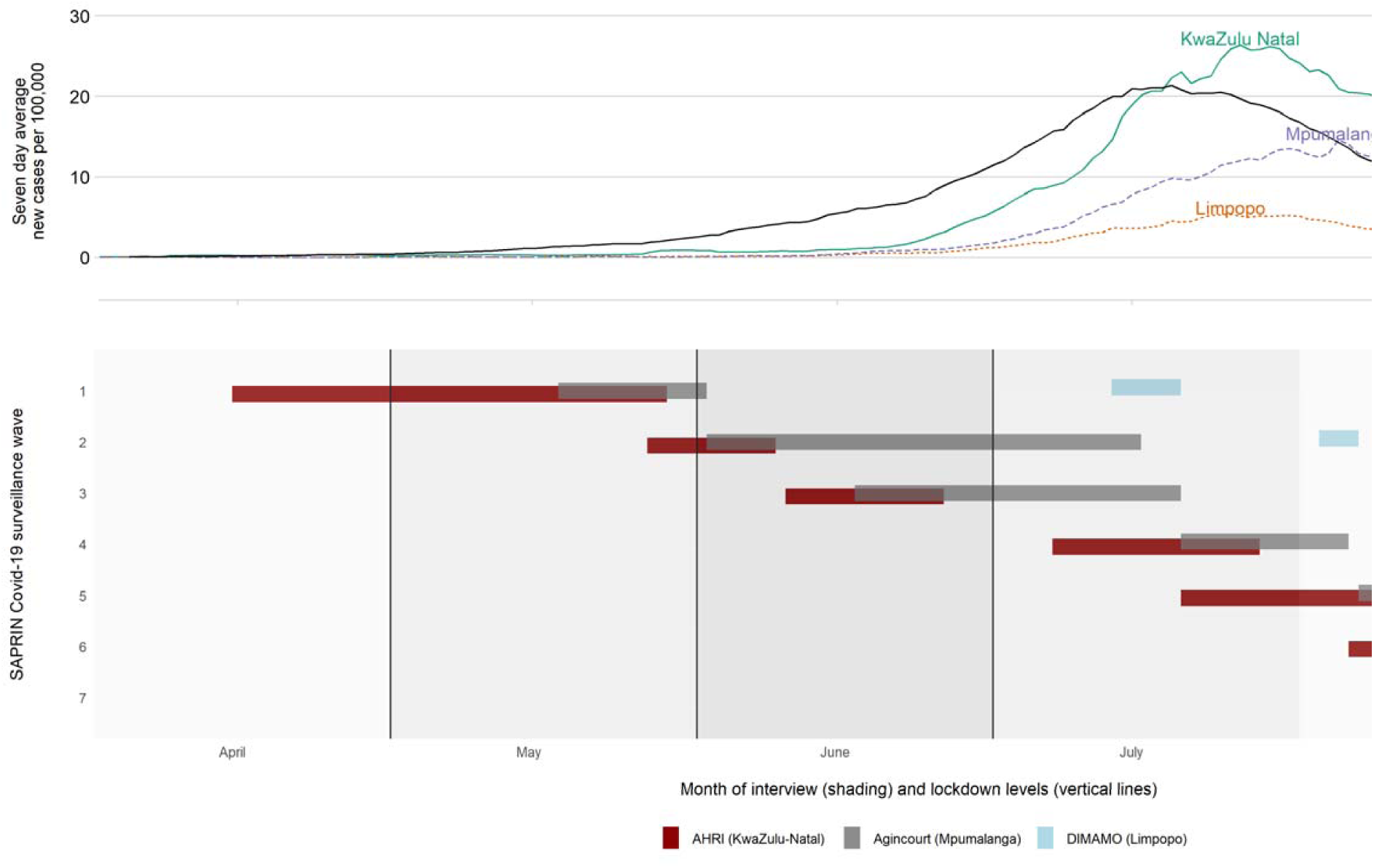
Epidemic curve and interview rounds across SAPRIN nodes.

Descriptive statistics for all sampled households with a valid telephone number are shown in Supplementary Table 1. In one-sixth (16.4%) of households no-one had completed secondary education, while in 14.7% a household member had completed a post-secondary qualification. Wealthier households were more likely to have a valid telephone number and more likely to participate at AHRI and Agincourt. Households were large (median of six resident and non-resident members, of whom two were under age 18). Households had a mean of 1.6 employed members, 38% of all households received one or more old-age pensions and 63% received any other government grant.

Figure 2 describes questionnaire response over time and by location. In terms of knowledge and behaviour, self-reported satisfaction with knowledge about Covid-19 rose over time at all three nodes, from around 48.3% in April and May to 84.5% in August. Daily visitors to the house were consistently low, peaking at 11.9% of households having one or more visitors in the previous day in May. The proportion of households with members going out increased over time, from 28.2% in April to a peak of 79.5% in June. There was, however, a notable drop at AHRI in KwaZulu-Natal, concurrent with reports of local Covid-19 transmission, from 76.2% in June to 31.7% in July and 18.0% in August. Facemask use rose rapidly to become almost universal at all three nodes by June.

**Figure 2:**
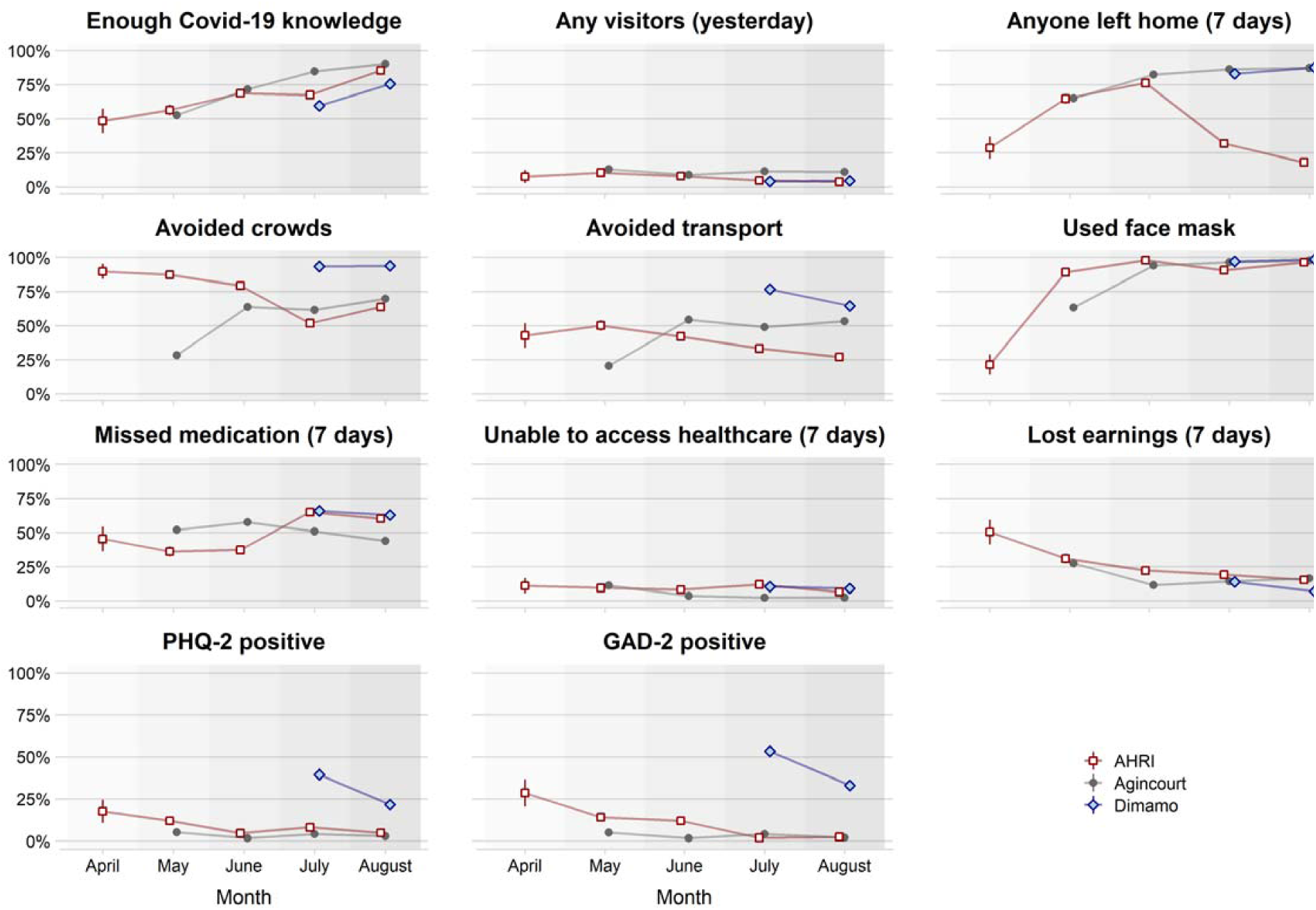
Knowledge, behaviour and impact of Covid-19 and related regulations at SAPRIN nodes. PHQ-2: Patient Health Questionnaire 2-item; GAD-2: Generalized Anxiety Disorder 2-item. Values are proportions and 95% confidence intervals of household primary respondents. Precise values in Supplementary Table 2.

Reported inability to access healthcare remained low and relatively stable over time. However, around half of households reported missing daily medication, and while levels were stable at Agincourt and DIMAMO, they almost doubled at AHRI from 37.4% in June to 65.0% as the epidemic arrived locally in July. The proportion of households reporting lost earnings due to Covid-19 regulations dropped substantially as lockdown was reduced from Level 5 to Level 4, and again when it was lowered to Level 3, remaining steady thereafter. Finally, the proportion of individuals screening positive for possible anxiety and depression fell over time at AHRI and DIMAMO and stayed low at Agincourt throughout the period.

After accounting for study node and month of interview, and despite variation in household composition, no household characteristics were substantively associated with self-reported satisfactory Covid-19 knowledge, residents having left the home in the past week, avoiding travel or facemask use (Table 2). Households with more educated individuals were more likely to report having had visitors the previous day and less likely to have avoided crowded spaces, perhaps reflecting urban location. Households with older and more pension-recipients were non-significantly more likely to have a recent unmet health need, but less likely to have been unable to access medicine (Table 3). These same two factors, as well as having a member who had completed secondary but not post-secondary education, all predicted less lost earnings. Finally, symptoms of depression and anxiety were non-significantly lower in households receiving pension grants.

**Table 1:** Response patterns for eligible households by SAPRIN node and interview round.

| **Node** | **Wave** | **Phone out of order**<br>**/wrong number** |  | **Phone not**<br>**answered** |  | **Refused**<br>**call** |  | **Successful** |  |
| --- | --- | --- | --- | --- | --- | --- | --- | --- | --- |
| Agincourt | 1 | 257 | (14.6%) | 190 | (10.8%) | 155 | (6.2%) | 1,159 | (65.8%) |
|  | 2 | 35 | (3.1%) | 46 | (4.1%) | 28 | (2.5%) | 1,013 | (90.3%) |
|  | 3 | 35 | (3.3%) | 48 | (4.6%) | 24 | (2.3%) | 944 | (89.8%) |
|  | 4 | 56 | (4.8%) | 72 | (6.2%) | 27 | (2.3%) | 1,004 | (86.6%) |
|  | 5 | 93 | (8.1%) | 76 | (6.6%) | 32 | (2.8%) | 951 | (82.6%) |
|  | 6 | 42 | (7.4%) | 63 | (11.1%) | 26 | (4.6%) | 439 | (77.0%) |
| AHRI | 1 | 69 | (9.2%) | 69 | (9.2%) | 8 | (1.1%) | 604 | (80.5%) |
|  | 2 | 63 | (8.4%) | 140 | (18.7%) | 17 | (2.3%) | 530 | (70.7%) |
|  | 3 | 49 | (6.5%) | 185 | (24.7%) | 24 | (3.2%) | 492 | (65.6%) |
|  | 4 | 69 | (9.2%) | 90 | (12.0%) | 20 | (2.7%) | 571 | (76.1%) |
|  | 5 | 79 | (10.5%) | 136 | (18.1%) | 18 | (2.4%) | 517 | (68.9%) |
|  | 6 | 52 | (6.9%) | 158 | (21.1%) | 8 | (1.1%) | 532 | (70.9%) |
|  | 7 | 45 | (7.3%) | 126 | (20.6%) | 9 | (1.5%) | 433 | (70.6%) |
| DIMAMO | 1 | 82 | (9.3%) | 63 | (7.2%) | 31 | (3.5%) | 705 | (80.0%) |
|  | 2 | 87 | (10.0%) | 78 | (8.9%) | 32 | (3.7%) | 677 | (77.5%) |
|  | 3 | 41 | (8.0%) | 41 | (8.0%) | 38 | (7.4%) | 395 | (76.7%) |
| Total |  | 1,053 | (7.6%) | 1,485 | (10.7%) | 435 | (3.1%) | 10,966 | (78.7%) |

All interviews up to 31<sup>st</sup> August 2020. Last round at each node is incomplete, hence lower total numbers. 464 of the 2,225 households initially sampled by Agincourt, and 119 of the 1,000 households initially sampled by DIMAMO did not have valid telephone numbers and are excluded here. From wave 2, Agincourt only sought to reach the 1159 households that participated in wave 1, although in waves 2 and 3 some households were unintentionally not contacted and in wave 5 seven households were intentionally not contacted due to either involvement in other intensive studies or having left the area.

**Table 2:** Knowledge and behaviour levels regarding Covid-19 and household characteristics.

| Node | Enough knowledge |  | Any visitors |  | Left home |  |
| --- | --- | --- | --- | --- | --- | --- |
| AHRI | 1.00 |  | 1.00 |  | 1.00 |  |
| Agincourt | 1.11 | [1.07 - 1.14] | 1.88 | [1.51 - 2.34] | 1.88 | [1.80 - 1.97] |
| DIMAMO | 0.87 | [0.81 - 0.93] | 0.78 | [0.49 - 1.23] | 2.18 | [2.05 - 2.33] |
| Month of interview |  |  |  |  |  |  |
| April | 0.94 | [0.77 - 1.14] | 0.99 | [0.52 - 1.90] | 0.66 | [0.49 - 0.88] |
| May | 1.00 |  | 1.00 |  | 1.00 |  |
| June | 1.32 | [1.25 - 1.39] | 0.72 | [0.59 - 0.87] | 1.27 | [1.22 - 1.33] |
| July | 1.44 | [1.37 - 1.51] | 0.73 | [0.62 - 0.87] | 1.00 | [0.95 - 1.04] |
| August | 1.65 | [1.58 - 1.73] | 0.67 | [0.56 - 0.81] | 0.91 | [0.87 - 0.96] |
| Household members (per person) |  |  |  |  |  |  |
| Children | 1.00 | [1.00 - 1.01] | 0.99 | [0.94 - 1.06] | 1.02 | [1.01 - 1.04] |
| Working-age adults | 1.00 | [0.99 - 1.00] | 0.99 | [0.95 - 1.04] | 1.01 | [1.00 - 1.02] |
| Pension-age adults | 0.98 | [0.95 - 1.00] | 1.16 | [0.97 - 1.41] | 0.99 | [0.96 - 1.03] |
| Education |  |  |  |  |  |  |
| Less than complete secondary | 1.00 |  | 1.00 |  | 1.00 |  |
| Complete secondary | 1.03 | [1.00 - 1.07] | 0.95 | [0.77 - 1.17] | 0.99 | [0.95 - 1.03] |
| Diploma/certificate/degree | 1.02 | [0.95 - 1.08] | 1.15 | [0.79 - 1.66] | 1.01 | [0.94 - 1.08] |
| Household income sources in 2019 |  |  |  |  |  |  |
| Full/part-time employees | 1.01 | [0.99 - 1.02] | 1.03 | [0.96 - 1.10] | 1.02 | [1.01 - 1.03] |
| Pension grants | 0.99 | [0.96 - 1.02] | 0.91 | [0.74 - 1.11] | 0.96 | [0.92 - 1.00] |
| Non-pension grants | 1.00 | [0.99 - 1.01] | 0.96 | [0.90 - 1.02] | 0.99 | [0.98 - 1.00] |
| Household asset index quintiles | 1.01 | [1.00 - 1.02] | 0.97 | [0.91 - 1.03] | 1.01 | [1.00 - 1.02] |
| N | 9,259 |  | 9,254 |  | 9,173 |  |

| Node | Avoid crowds |  | Avoid transport |  | Use facemask |  |
| --- | --- | --- | --- | --- | --- | --- |
| AHRI | 1.00 |  | 1.00 |  | 1.00 |  |
| Agincourt | 0.82 | [0.79 - 0.85] | 1.20 | [1.13 - 1.27] | 0.96 | [0.94 - 0.97] |
| DIMAMO | 1.34 | [1.29 - 1.41] | 1.84 | [1.68 - 2.01] | 0.99 | [0.97 - 1.01] |
| Month of interview |  |  |  |  |  |  |
| April | 1.57 | [1.44 - 1.70] | 1.53 | [1.22 - 1.92] | 0.28 | [0.19 - 0.40] |
| May | 1.00 |  | 1.00 |  | 1.00 |  |
| June | 1.36 | [1.29 - 1.44] | 1.56 | [1.43 - 1.70] | 1.30 | [1.26 - 1.34] |
| July | 1.14 | [1.08 - 1.21] | 1.37 | [1.26 - 1.49] | 1.28 | [1.24 - 1.32] |
| August | 1.29 | [1.22 - 1.37] | 1.33 | [1.22 - 1.45] | 1.32 | [1.28 - 1.36] |
| Household members (per person) |  |  |  |  |  |  |
| Children | 1.00 | [0.99 - 1.01] | 1.00 | [0.98 - 1.02] | 1.00 | [1.00 - 1.00] |
| Working-age adults | 1.01 | [1.01 - 1.02] | 1.03 | [1.01 - 1.04] | 1.00 | [1.00 - 1.00] |
| Pension-age adults | 1.04 | [1.01 - 1.07] | 1.03 | [0.97 - 1.09] | 1.00 | [0.99 - 1.01] |
| Education |  |  |  |  |  |  |
| Less than complete secondary | 1.00 |  | 1.00 |  | 1.00 |  |
| Complete secondary | 0.99 | [0.95 - 1.03] | 0.99 | [0.94 - 1.06] | 1.00 | [0.99 - 1.02] |
| Diploma/certificate/degree | 0.94 | [0.89 - 1.00] | 1.03 | [0.93 - 1.14] | 1.00 | [0.97 - 1.02] |
| Household income sources in 2019 |  |  |  |  |  |  |
| Full/part-time employees | 0.99 | [0.98 - 1.00] | 0.98 | [0.96 - 1.00] | 1.00 | [1.00 - 1.00] |
| Pension grants | 0.97 | [0.94 - 1.01] | 0.99 | [0.93 - 1.05] | 1.00 | [0.98 - 1.02] |
| Non-pension grants | 1.00 | [0.99 - 1.01] | 1.01 | [0.99 - 1.02] | 1.00 | [0.99 - 1.00] |
| Household asset index quintiles | 1.00 | [0.99 - 1.02] | 1.00 | [0.99 - 1.02] | 1.00 | [1.00 - 1.00] |
| N | 9,184 |  | 9,184 |  | 9,184 |  |

**Table 3:**
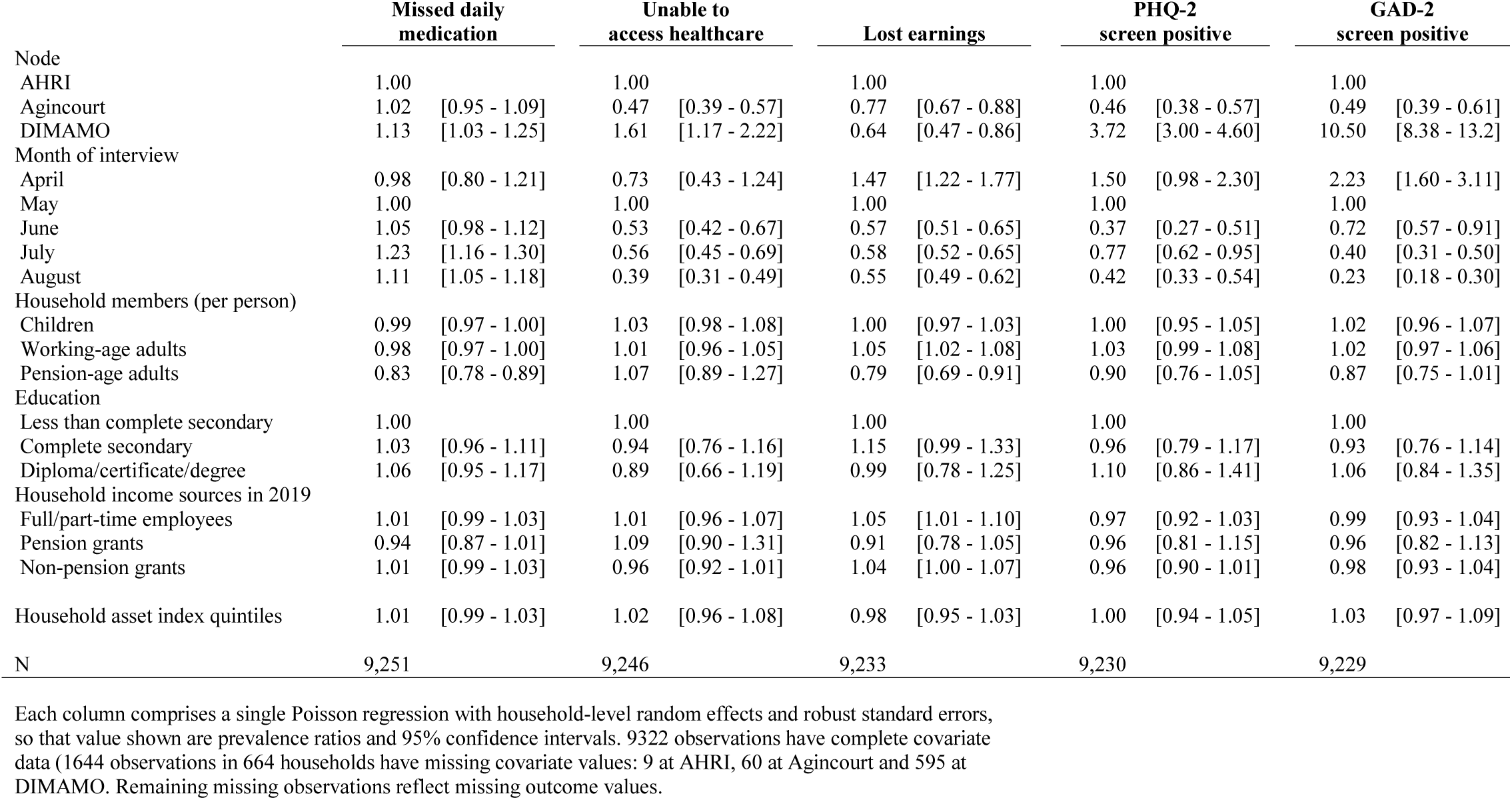
Social, economic and health impacts of Covid-19 and household characteristics.

|  | **Missed daily**<br>**medication** |  | **Unable to**<br>**access healthcare** |  | **Lost earnings** |  | **PHQ-2**<br>**screen positive** |  | **GAD-2**<br>**screen positive** |  |
| --- | --- | --- | --- | --- | --- | --- | --- | --- | --- | --- |
| Node |  |  |  |  |  |  |  |  |  |  |
| AHRI | 1.00 |  | 1.00 |  | 1.00 |  | 1.00 |  | 1.00 |  |
| Agincourt | 1.02 | [0.95 - 1.09] | 0.47 | [0.39 - 0.57] | 0.77 | [0.67 - 0.88] | 0.46 | [0.38 - 0.57] | 0.49 | [0.39 - 0.61] |
| DIMAMO | 1.13 | [1.03 - 1.25] | 1.61 | [1.17 - 2.22] | 0.64 | [0.47 - 0.86] | 3.72 | [3.00 - 4.60] | 10.50 | [8.38 - 13.2] |
| Month of interview |  |  |  |  |  |  |  |  |  |  |
| April | 0.98 | [0.80 - 1.21] | 0.73 | [0.43 - 1.24] | 1.47 | [1.22 - 1.77] | 1.50 | [0.98 - 2.30] | 2.23 | [1.60 - 3.11] |
| May | 1.00 |  | 1.00 |  | 1.00 |  | 1.00 |  | 1.00 |  |
| June | 1.05 | [0.98 - 1.12] | 0.53 | [0.42 - 0.67] | 0.57 | [0.51 - 0.65] | 0.37 | [0.27 - 0.51] | 0.72 | [0.57 - 0.91] |
| July | 1.23 | [1.16 - 1.30] | 0.56 | [0.45 - 0.69] | 0.58 | [0.52 - 0.65] | 0.77 | [0.62 - 0.95] | 0.40 | [0.31 - 0.50] |
| August | 1.11 | [1.05 - 1.18] | 0.39 | [0.31 - 0.49] | 0.55 | [0.49 - 0.62] | 0.42 | [0.33 - 0.54] | 0.23 | [0.18 - 0.30] |
| Household members (per person) |  |  |  |  |  |  |  |  |  |  |
| Children | 0.99 | [0.97 - 1.00] | 1.03 | [0.98 - 1.08] | 1.00 | [0.97 - 1.03] | 1.00 | [0.95 - 1.05] | 1.02 | [0.96 - 1.07] |
| Working-age adults | 0.98 | [0.97 - 1.00] | 1.01 | [0.96 - 1.05] | 1.05 | [1.02 - 1.08] | 1.03 | [0.99 - 1.08] | 1.02 | [0.97 - 1.06] |
| Pension-age adults | 0.83 | [0.78 - 0.89] | 1.07 | [0.89 - 1.27] | 0.79 | [0.69 - 0.91] | 0.90 | [0.76 - 1.05] | 0.87 | [0.75 - 1.01] |
| Education |  |  |  |  |  |  |  |  |  |  |
| Less than complete secondary | 1.00 |  | 1.00 |  | 1.00 |  | 1.00 |  | 1.00 |  |
| Complete secondary | 1.03 | [0.96 - 1.11] | 0.94 | [0.76 - 1.16] | 1.15 | [0.99 - 1.33] | 0.96 | [0.79 - 1.17] | 0.93 | [0.76 - 1.14] |
| Diploma/certificate/degree | 1.06 | [0.95 - 1.17] | 0.89 | [0.66 - 1.19] | 0.99 | [0.78 - 1.25] | 1.10 | [0.86 - 1.41] | 1.06 | [0.84 - 1.35] |
| Household income sources in 2019 |  |  |  |  |  |  |  |  |  |  |
| Full/part-time employees | 1.01 | [0.99 - 1.03] | 1.01 | [0.96 - 1.07] | 1.05 | [1.01 - 1.10] | 0.97 | [0.92 - 1.03] | 0.99 | [0.93 - 1.04] |
| Pension grants | 0.94 | [0.87 - 1.01] | 1.09 | [0.90 - 1.31] | 0.91 | [0.78 - 1.05] | 0.96 | [0.81 - 1.15] | 0.96 | [0.82 - 1.13] |
| Non-pension grants | 1.01 | [0.99 - 1.03] | 0.96 | [0.92 - 1.01] | 1.04 | [1.00 - 1.07] | 0.96 | [0.90 - 1.01] | 0.98 | [0.93 - 1.04] |
| Household asset index quintiles | 1.01 | [0.99 - 1.03] | 1.02 | [0.96 - 1.08] | 0.98 | [0.95 - 1.03] | 1.00 | [0.94 - 1.05] | 1.03 | [0.97 - 1.09] |
| N | 9,251 |  | 9,246 |  | 9,233 |  | 9,230 |  | 9,229 |  |

Each column comprises a single Poisson regression with household-level random effects and robust standard errors, so that value shown are prevalence ratios and 95% confidence intervals. 9322 observations have complete covariate data (1644 observations in 664 households have missing covariate values: 9 at AHRI, 60 at Agincourt and 595 at DIMAMO. Remaining missing observations reflect missing outcome values.

## DISCUSSION

Using rapid, repeated, telephonic interviews across three provinces in South Africa, we observed how households in rural and peri-urban areas responded to, and were affected by, national NPIs enacted to minimize the epidemic spread of Covid-19. As both NPIs and the epidemic spread across the country, our longitudinal surveillance programme captured the impact of both processes.

Our first key finding was that the national public health measures and messaging were effective in several ways. Respondents across three provinces showed consistent improvements in satisfaction with their understanding of the epidemic. There was early concern that South Africa’s public health messaging was insufficiently contextualized to the country’s varying social and economic conditions. However, both politicians and scientists conducted televised national press conferences, supported by provincial and local follow-up. While causality cannot be proven, in line with evidence elsewhere [23], our respondents reported rapid and comprehensive compliance with communicated messages, including facemask use, and active avoidance of crowds and transport. Several of these behaviours remained prevalent even as formal lockdown NPIs were relaxed.

Second, we identified substantial behaviour change as Covid-19 arrived locally. This was particularly noticeable in KwaZulu-Natal, where rapid epidemic expansion in the local district in early July coincided with rapid decline in household members leaving home and concomitant increases in missed daily medications and inability to access needed healthcare. This response reflected local epidemic dynamics – the national epidemic started in the Western Cape, extending first to the adjacent Eastern Cape, then to densely populated Gauteng and eThekwini in KwaZulu-Natal, before expanding to more the rural east and north of South Africa captured in this study. The impact of regulations and epidemic trajectories on travel is particularly pertinent in our study settings: medium- and long-distance circular labour migration to urban areas is highly prevalent and vital to economic wellbeing for rural and peri-urban South African households [24]. Bans on long-distance travel had potentially substantial economic implications for people not able to return to work, although such travel bans might also explain in part the limited epidemics seen in these rural areas even as NPIs were relaxed.

Third, mental health findings are reassuring. At all three nodes we observed declining depression and anxiety symptoms over time, based on validated screening scales. Symptom frequency was notably higher at the DIMAMO peri-urban node, perhaps reflecting concern of greater risk arising from the nearby city of Polokwane. Encouragingly, even when Covid-19 arrived locally at AHRI, depression and anxiety rates did not increase. Comparisons are complex, but our findings align with national South African data suggesting lower Covid-19 mental health impact in poorer and more rural areas [23]. Longitudinal surveillance across a range of settings using harmonised measures will help unpick how much mental health is affected directly by Covid-19 disease fears, and indirectly by secondary social and economic effects.

Our analysis raises concern about unmet need for healthcare. Almost half of households reported that members had recently missed daily medication doses, and 5-10% of households reported that a member wanted but been unable to access healthcare in the past seven days, levels similar to other South African surveys [13]. Notably, epidemic arrival at AHRI had diverging effects: unmet healthcare need changed little, but missed medication rates almost doubled. Such patterns suggest that households may be calculating trade-offs between Covid and non-Covid risks, being willing to risk physical proximity to attend clinic [25], but not collect medicine [26]. Additional information is needed to determine whether unmet healthcare needs represent operational, mobility, transport costs/availability or other issues – and to what extent they were the result of or exacerbated by the Covid-19 epidemic or regulations. Data covering pre- and post-Covid-19 would help unpick these effects, as would qualitative investigation of household decision-making during lockdown.

Finally, we found that households with greater numbers of older members and pension-recipients reported greater unmet healthcare needs, but were less affected by lost earnings and had fewer mental health concerns. South African non-contributory pensions – broad national government support schemes that are often the largest household income source in these relatively rural settings with very high unemployment rates – have previously been linked to positive physical and mental health outcomes [27, 28]. Our study suggests that such government support structures likely play an essential role maintaining household security in crisis contexts such as Covid-19, providing a guaranteed income to vulnerable populations. The government’s temporary supplementation of grant programmes early in the epidemic may have additionally helped [29], however it will be important to observe if the ending of supplementation in October 2020 reverses effects.

This analysis presents an overview of key insights across time from multiple sites across South Africa. However, there are several follow-on analyses to consider that would further contextualize our findings. First, data can be analysed longitudinally at the household or individual level to evaluate trajectories of behaviour and impact as the COVID-19 epidemic continues to play out in rural and peri-urban environments. For example, it will be important to evaluate the impact of new government policies such as the ending of temporary increases in non-contributory grants. Second, these behavioural data can be linked to experience of Covid-19 symptoms and individual and population health outcomes to evaluate how risk perceptions and reactions were associated with health outcomes. Third, more in-depth analysis of how households’ historical and current age, gender, employment and migration composition, as well as pre-existing co-morbidities, affects current Covid-19 and NPI impact will help identify those most in need of support during such crises. The ongoing SAPRIN Covid-19 surveillance programme will allow measurement of these factors longitudinally throughout the epidemic’s course.

### Strengths and limitations

This study has limitations. As with all observational studies, generalizability beyond our study population, in this case rural and peri-urban areas of eastern South Africa, is uncertain. This concern is tempered by our ability to compare and combine data across multiple sites and to triangulate with other studies of Covid-19 impact in South Africa and sub-Saharan African. While household cellphone ownership is high, there was evidence that poorer households in these areas were somewhat less likely to participate, which will affect response levels although less so the trends we focus on. Our data are self-reported, and thus represent perceived needs and impacts, and reported behaviour change may reflect desirability biases. However, even with such biases, findings provide insight into the perceptions and lived experiences of these communities. Again, triangulating our findings with digital data sources can help. Finally, we do not have data on identical questions from the pre-Covid period, however, we are able to leverage similar information on many topics from earlier surveillance.

Nevertheless, the study also has several strengths, notably a clearly defined sampling base, high response and low attrition rates, frequent follow-up and linkage to pre-Covid household data. Many of these benefits arise from the nature of the existing SAPRIN surveillance infrastructure, reinforcing the importance of long-term population-based surveillance systems that collect social, demographic and health data. This work demonstrates that surveillance systems can be rapidly repurposed to answer emergency health needs, including: (i) rapid pathogen data acquisition; (ii) identification of susceptible populations; (iii) assessment of behavioural and biomedical interventions; and (iv) development of mitigation strategies [30].

SAPRIN nodes have been working with their local communities for 20-28 years. Such long-term engagement ensures deep understanding and community buy-in allowing rapid implementation and sustained high-intensity follow-up with minimal dropout. The network nature of SAPRIN also allowed each node to flexibly implement an overarching protocol. Furthermore, use of telephonic call centres at each node enabled rapid survey roll-out based on previously provided informed consent for personal calls and substantially reduced Covid-19 infection risk for study staff and research participants. Since these call centres employ locally recruited staff, we were able to reach population segments that online surveys (in a country with limited access to internet in rural areas), and even random-digit dialling approaches (in a country with 11 official spoken languages and numerous dialects) struggle to capture. Additionally, we could link self-response survey data to other data sources within the SAPRIN databases. These include the previously collected sociodemographic data used in this study and biological samples collected as part of the Covid-19 surveillance project. SAPRIN can also link to data on public sector healthcare utilization and laboratory test results through memoranda of understanding with government departments. SAPRIN’s ongoing expansion will also allow comparisons to be made in the future to well-characterised urban sites.

### Conclusion

South Africans in three rural and peri-urban areas were to a great extent willing and able to comply with national government regulations and recommendations on social interaction and other risk behaviours relating to Covid-19, despite limited resources and substantial economic need to travel. This rapid uptake of preventative behaviours reflects clear government messaging and population willingness to comply even in settings where enforcement measures were limited. The arrival of the epidemic locally, even as official NPIs were relaxed, led to further self-imposed behavioural restrictions, some of which led to difficulties with healthcare access. However, the economic and mental health effects of NPIs continued to decline as the measures were eased. Our findings highlight the importance of monitoring the possible deleterious secondary impacts of NPIs in epidemic conditions. Our results reinforce the principle that NPIs should be adjusted based on epidemic cycle and that mitigating measures will be required against anticipated and unanticipated secondary impacts. All these factors should be considered when setting, adjusting and relaxing NPIs in lower-income settings, especially as urgently established national policies give way to differentiated, decentralised approaches across diverse subnational environments.

## Data Availability

Data collected in the SAPRIN Covid-19 high-intensity surveillance study will be made available in pseudonymized form through Figshare and the SAPRIN data repository (http://saprindata.samrc.ac.za/index.php/catalog). Data reported in this article will be available on peer-reviewed publication. Data from ongoing SAPRIN surveillance is shared on the SAPRIN data repository.

## List of abbreviations

AHRI: Africa Health Research Institute
GAD-2: Generalized Anxiety Disorder 2-item
HDSS: Health and Demographic Surveillance System
NPI: Non-Pharmaceutical Intervention
PHQ-2: Patient Health Questionnaire 2-item
REC: Research Ethics Committee
SAPRIN: South African Population Research Infrastructure Network

## Consent for publication

Not applicable

## Availability of data and materials

The datasets reported in this article will be available on publication through the SAPRIN data repository (http://saprindata.samrc.ac.za/index.php/catalog) in pseudonymized form.

## Competing interests

The authors declare that they have no competing interests.

## Funding

Core surveillance activities including the Covid-19 telephonic surveillance at all three nodes are wholly or in part funded by the South African Department of Science and Innovation through the South African Population Research Infrastructure Network hosted by the South African Medical Research Council.

The MRC/Wits Rural Public Health and Health Transitions Research Unit which hosts the Agincourt Health and Socio-Demographic Surveillance System is also supported by the University of the Witwatersrand and Medical Research Council, South Africa. DIMAMO Population Health Research Centre receives funding from the National Institutes of Health and is supported by the Human Heredity and Health in Africa (H3Africa) Consortium. African Health Research Institute receives funding from the Wellcome Trust (Grant 201433/Z/16/Z) for aspects of its health and demographic surveillance.

GH is supported by a fellowship from the Royal Society and the Wellcome Trust (210479/Z/18/Z). EBW is supported by the National Institutes of Health (NIAID K08AI118538 and FIC R21TW011687).

The funders had no role in study design, data collection and analysis, decision to publish, or preparation of the manuscript.

## Authors’ contributions

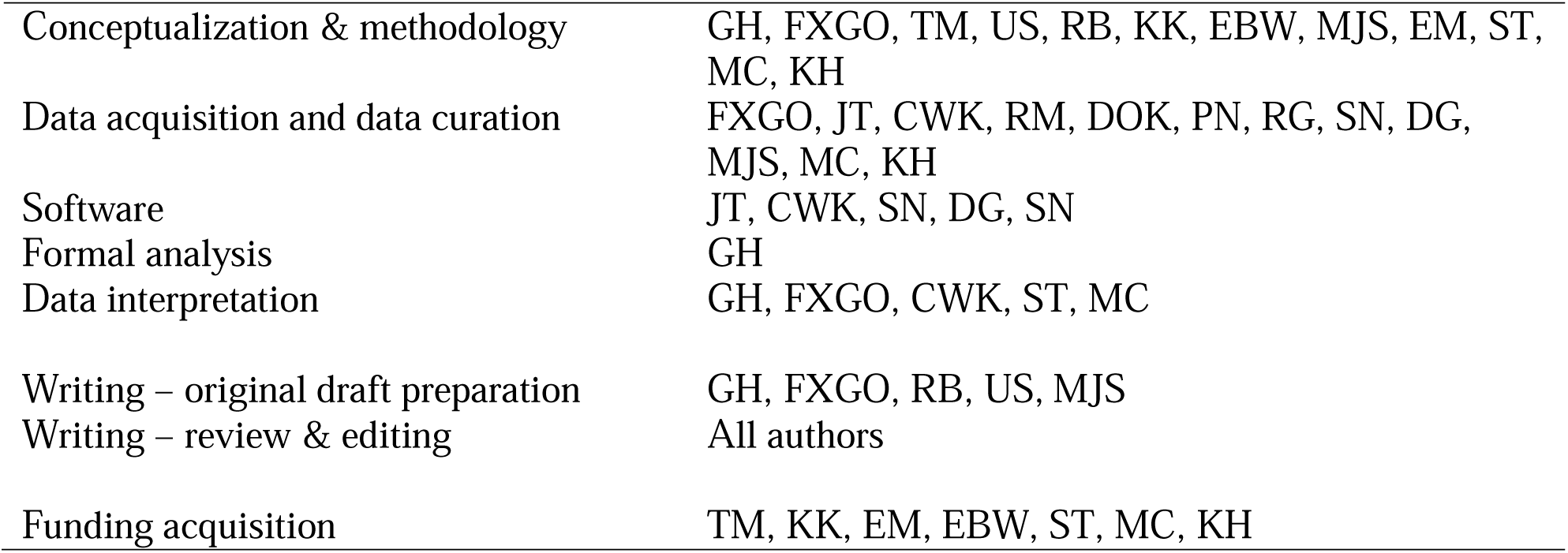

All authors contributed to critical revision of the work, final revisions to the text and agree to be accountable for the work.

## Acknowledgements

We gratefully acknowledge the enormous efforts of all the call-centre agents at the three study sites who worked intensively throughout, and the participants who responded multiple times despite difficult social and economic conditions.

## SUPPLEMENTARY MATERIAL

**Supplementary Figure 1:**
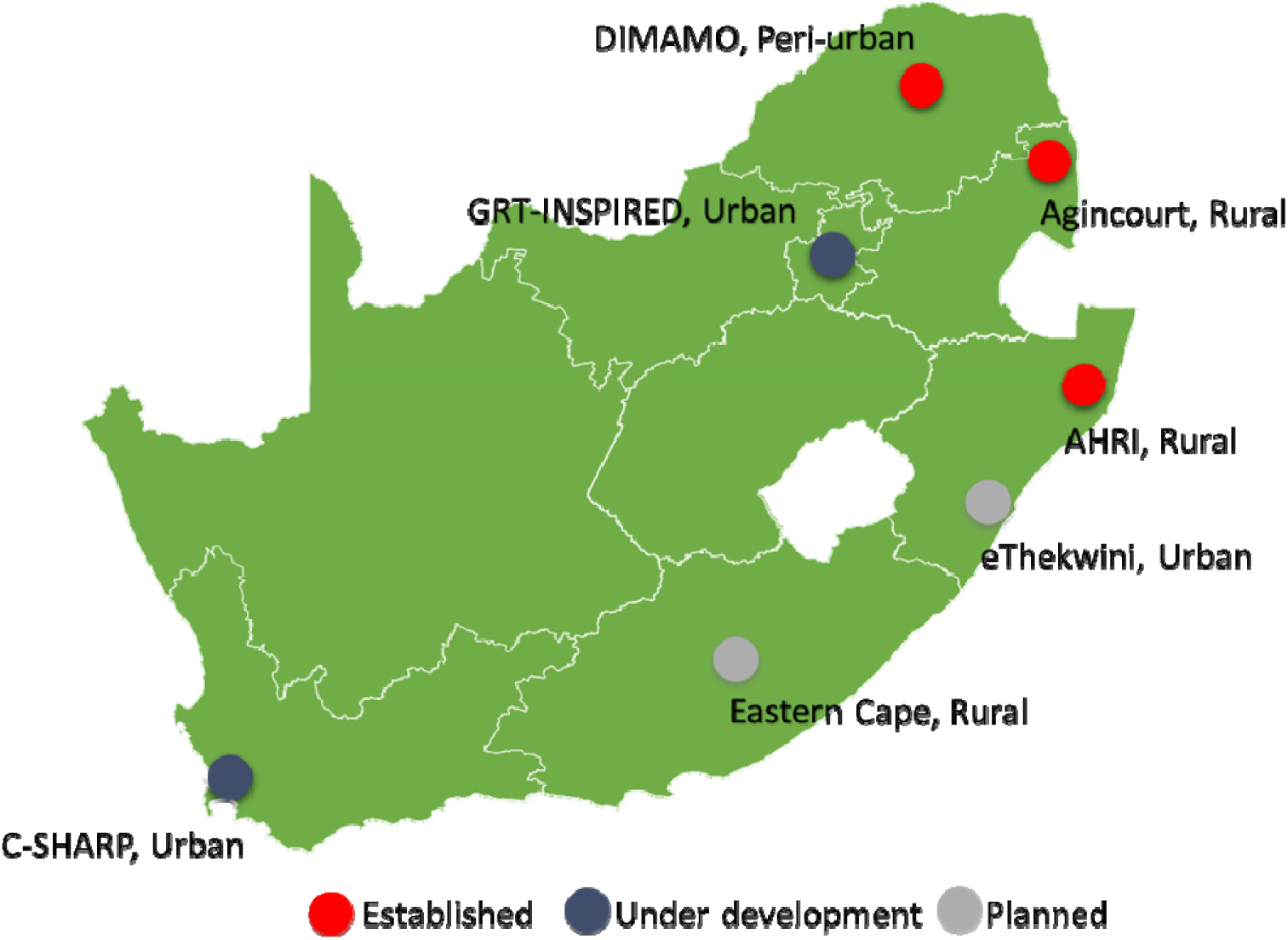
Map of SAPRIN nodes as of September 2020.

**Supplementary Table 1:**
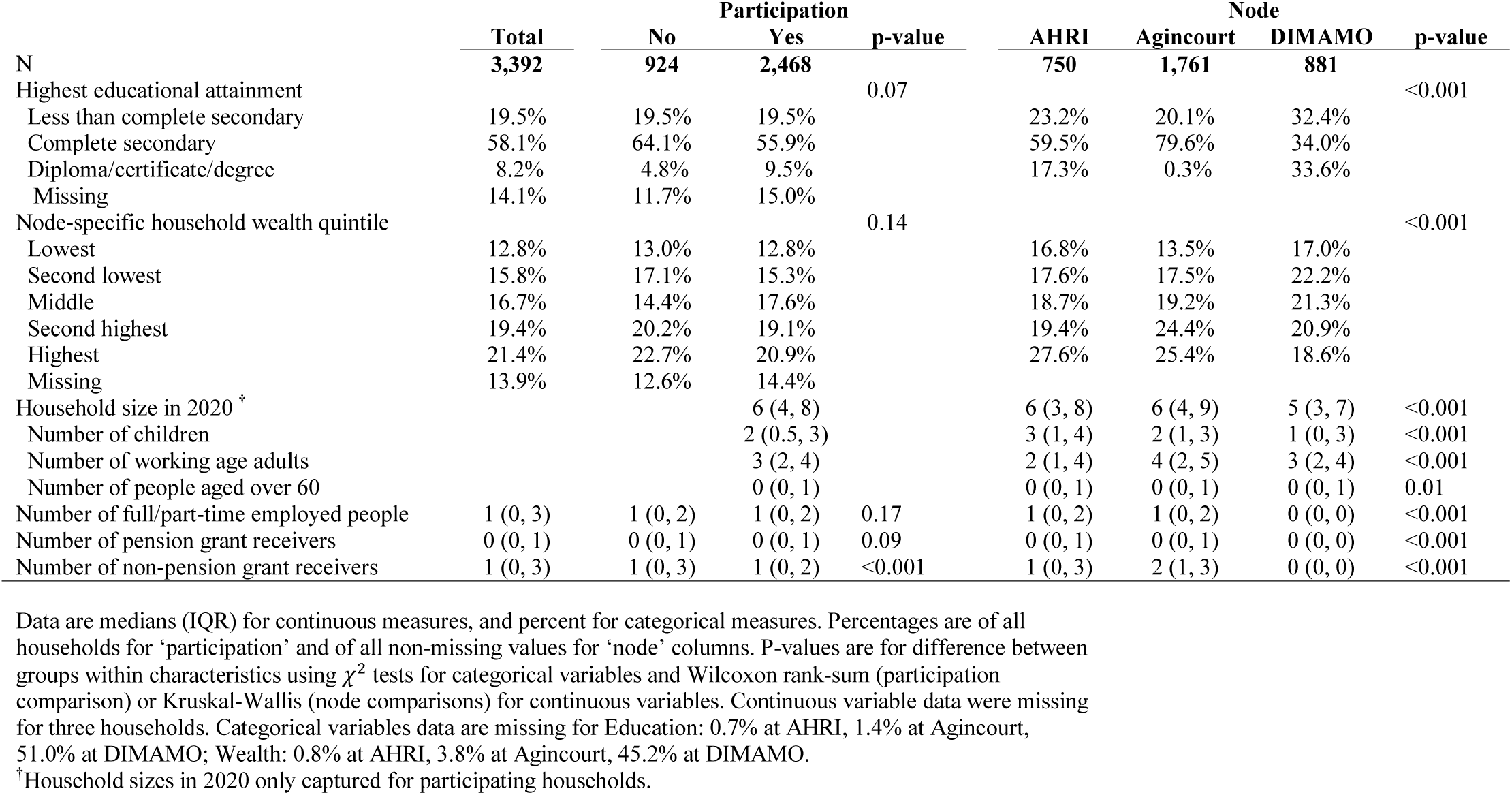
Descriptive statistics for eligible households.

|  | Participation |  |  |  | Node |  |  | p-value |
| --- | --- | --- | --- | --- | --- | --- | --- | --- |
|  | Total | No | Yes | p-value | AHRI | Agincourt | DIMAMO |  |
| N | 3,392 | 924 | 2,468 |  | 750 | 1,761 | 881 |  |
| Highest educational attainment |  |  |  | 0.07 |  |  |  | <0.001 |
| Less than complete secondary | 19.5% | 19.5% | 19.5% |  | 23.2% | 20.1% | 32.4% |  |
| Complete secondary | 58.1% | 64.1% | 55.9% |  | 59.5% | 79.6% | 34.0% |  |
| Diploma/certificate/degree | 8.2% | 4.8% | 9.5% |  | 17.3% | 0.3% | 33.6% |  |
| Missing | 14.1% | 11.7% | 15.0% |  |  |  |  |  |
| Node-specific household wealth quintile |  |  |  | 0.14 |  |  |  | <0.001 |
| Lowest | 12.8% | 13.0% | 12.8% |  | 16.8% | 13.5% | 17.0% |  |
| Second lowest | 15.8% | 17.1% | 15.3% |  | 17.6% | 17.5% | 22.2% |  |
| Middle | 16.7% | 14.4% | 17.6% |  | 18.7% | 19.2% | 21.3% |  |
| Second highest | 19.4% | 20.2% | 19.1% |  | 19.4% | 24.4% | 20.9% |  |
| Highest | 21.4% | 22.7% | 20.9% |  | 27.6% | 25.4% | 18.6% |  |
| Missing | 13.9% | 12.6% | 14.4% |  |  |  |  |  |
| Household size in 2020 <sup>†</sup> |  |  | 6 (4, 8) |  | 6 (3, 8) | 6 (4, 9) | 5 (3, 7) | <0.001 |
| Number of children |  |  | 2 (0.5, 3) |  | 3 (1, 4) | 2 (1, 3) | 1 (0, 3) | <0.001 |
| Number of working age adults |  |  | 3 (2, 4) |  | 2 (1, 4) | 4 (2, 5) | 3 (2, 4) | <0.001 |
| Number of people aged over 60 |  |  | 0 (0, 1) |  | 0 (0, 1) | 0 (0, 1) | 0 (0, 1) | 0.01 |
| Number of full/part-time employed people | 1 (0, 3) | 1 (0, 2) | 1 (0, 2) | 0.17 | 1 (0, 2) | 1 (0, 2) | 0 (0, 0) | <0.001 |
| Number of pension grant receivers | 0 (0, 1) | 0 (0, 1) | 0 (0, 1) | 0.09 | 0 (0, 1) | 0 (0, 1) | 0 (0, 0) | <0.001 |
| Number of non-pension grant receivers | 1 (0, 3) | 1 (0, 3) | 1 (0, 2) | <0.001 | 1 (0, 3) | 2 (1, 3) | 0 (0, 0) | <0.001 |

Data are medians (IQR) for continuous measures, and percent for categorical measures. Percentages are of all households for 'participation' and of all non-missing values for 'node' columns. P-values are for difference between groups within characteristics using $\chi^2$ tests for categorical variables and Wilcoxon rank-sum (participation comparison) or Kruskal-Wallis (node comparisons) for continuous variables. Continuous variable data were missing for three households. Categorical variables data are missing for Education: 0.7% at AHRI, 1.4% at Agincourt, 51.0% at DIMAMO; Wealth: 0.8% at AHRI, 3.8% at Agincourt, 45.2% at DIMAMO.
<sup>†</sup>Household sizes in 2020 only captured for participating households.

**Supplementary Table 2:** Proportions of households affirming questions by node and month of interview.

**AHRI**
|  | **April** |  | **May** |  | **June** |  | **July** |  | **August** |  |
| --- | --- | --- | --- | --- | --- | --- | --- | --- | --- | --- |
| Enough knowledge | 48.3 | [39.3, 57.3] | 56.3 | [52.6, 60.1] | 68.6 | [65.5, 71.7] | 67.4 | [64.5, 70.4] | 85.3 | [83.2, 87.4] |
| Any visitors | 7.6 | [2.8, 12.4] | 10.4 | [8.1, 12.7] | 7.8 | [6.0, 9.6] | 4.7 | [3.4, 6.0] | 3.7 | [2.6, 4.9] |
| Left home | 28.2 | [20.1, 36.4] | 64.7 | [61.0, 68.3] | 76.2 | [73.3, 79.0] | 31.7 | [28.8, 34.6] | 18.0 | [15.7, 20.3] |
| Avoid crowds | 88.8 | [83.1, 94.5] | 87.0 | [84.5, 89.6] | 79.2 | [76.5, 82.0] | 51.8 | [48.6, 55.0] | 63.7 | [60.8, 66.6] |
| Avoid transport | 42.2 | [33.3, 51.2] | 49.9 | [46.1, 53.7] | 42.0 | [38.7, 45.3] | 33.2 | [30.2, 36.2] | 27.1 | [24.4, 29.8] |
| Use facemask | 20.7 | [13.3, 28.1] | 89.1 | [86.8, 91.5] | 98.0 | [97.0, 98.9] | 90.7 | [88.8, 92.5] | 96.5 | [95.3, 97.6] |
| Unable access medication | 45.8 | [36.8, 54.8] | 36.1 | [32.5, 39.8] | 37.4 | [34.2, 40.7] | 65.0 | [62.0, 68.0] | 60.1 | [57.2, 63.1] |
| Unmet healthcare need | 11.0 | [5.4, 16.7] | 9.3 | [7.1, 11.6] | 8.2 | [6.3, 10.0] | 11.9 | [9.9, 13.9] | 6.4 | [4.9, 7.8] |
| Lost earnings | 50.0 | [41.0, 59.0] | 30.9 | [27.4, 34.4] | 22.3 | [19.5, 25.1] | 19.1 | [16.7, 21.6] | 15.2 | [13.1, 17.4] |
| PHQ-2 | 16.9 | [10.2, 23.7] | 11.9 | [9.4, 14.4] | 4.6 | [3.2, 6.0] | 8.0 | [6.3, 9.7] | 4.8 | [3.5, 6.0] |
| GAD-2 | 28.0 | [19.9, 36.1] | 14.0 | [11.4, 16.6] | 12.1 | [9.9, 14.3] | 1.8 | [1.0, 2.7] | 2.3 | [1.4, 3.2] |

**Agincourt**
|  | **April** |  | **May** |  | **June** |  | **July** |  | **August** |  |
| --- | --- | --- | --- | --- | --- | --- | --- | --- | --- | --- |
| Enough knowledge |  |  | 52.5 | [49.5, 55.6] | 71.4 | [68.6, 74.2] | 84.6 | [83.0, 86.3] | 90.1 | [88.6, 91.6] |
| Any visitors |  |  | 12.9 | [10.9, 15.0] | 8.8 | [7.1, 10.6] | 11.3 | [9.9, 12.8] | 11.0 | [9.5, 12.6] |
| Left home |  |  | 65.0 | [62.1, 67.9] | 82.3 | [79.9, 84.7] | 86.3 | [84.7, 87.9] | 87.1 | [85.4, 88.8] |
| Avoid crowds |  |  | 28.3 | [25.6, 31.1] | 63.6 | [60.6, 66.5] | 61.0 | [58.8, 63.2] | 69.7 | [67.4, 71.9] |
| Avoid transport |  |  | 20.5 | [18.0, 23.0] | 54.0 | [51.0, 57.1] | 48.7 | [46.4, 51.0] | 53.4 | [51.0, 55.9] |
| Use facemask |  |  | 63.4 | [60.5, 66.4] | 93.8 | [92.3, 95.2] | 95.5 | [94.6, 96.5] | 97.7 | [96.9, 98.4] |
| Unable access medication |  |  | 51.9 | [48.9, 55.0] | 57.1 | [54.0, 60.1] | 51.3 | [49.0, 53.5] | 44.4 | [41.9, 46.9] |
| Unmet healthcare need |  |  | 11.2 | [9.3, 13.1] | 3.5 | [2.4, 4.7] | 2.4 | [1.7, 3.1] | 2.4 | [1.6, 3.1] |
| Lost earnings |  |  | 27.5 | [24.8, 30.2] | 11.4 | [9.4, 13.3] | 14.4 | [12.8, 16.0] | 16.3 | [14.5, 18.1] |
| PHQ-2 |  |  | 5.2 | [3.8, 6.5] | 1.6 | [0.8, 2.4] | 4.0 | [3.1, 4.9] | 2.8 | [2.0, 3.6] |
| GAD-2 |  |  | 5.0 | [3.6, 6.3] | 1.5 | [0.7, 2.2] | 4.0 | [3.1, 4.9] | 1.9 | [1.2, 2.5] |

**DIMAMO**
|  | **April** |  | **May** |  | **June** |  | **July** |  | **August** |  |
| --- | --- | --- | --- | --- | --- | --- | --- | --- | --- | --- |
| Enough knowledge |  |  |  |  |  |  | 59.6 | [56.0, 63.2] | 75.5 | [72.9, 78.0] |
| Any visitors |  |  |  |  |  |  | 4.0 | [2.5, 5.4] | 4.5 | [3.2, 5.7] |
| Left home |  |  |  |  |  |  | 83.0 | [80.2, 85.7] | 87.6 | [85.6, 89.6] |
| Avoid crowds |  |  |  |  |  |  | 93.3 | [91.4, 95.2] | 93.8 | [92.3, 95.3] |
| Avoid transport |  |  |  |  |  |  | 76.8 | [73.6, 80.0] | 64.4 | [61.5, 67.3] |
| Use facemask |  |  |  |  |  |  | 97.0 | [95.7, 98.3] | 98.6 | [97.9, 99.3] |
| Unable access medication |  |  |  |  |  |  | 66.0 | [62.5, 69.5] | 62.9 | [60.0, 65.8] |
| Unmet healthcare need |  |  |  |  |  |  | 10.2 | [8.0, 12.4] | 9.0 | [7.3, 10.8] |
| Lost earnings |  |  |  |  |  |  | 13.8 | [11.2, 16.3] | 6.9 | [5.4, 8.4] |
| PHQ-2 |  |  |  |  |  |  | 39.6 | [36.0, 43.2] | 21.6 | [19.2, 24.1] |
| GAD-2 |  |  |  |  |  |  | 53.5 | [49.8, 57.2] | 33.0 | [30.2, 35.8] |

**Total**
|  | **April** |  | **May** |  | **June** |  | **July** |  | **August** |  |
| --- | --- | --- | --- | --- | --- | --- | --- | --- | --- | --- |
| Enough knowledge | 48.3 | [39.3, 57.3] | 54.0 | [51.6, 56.4] | 70.1 | [68.0, 72.2] | 74.8 | [73.4, 76.2] | 84.5 | [83.3, 85.7] |
| Any visitors | 7.6 | [2.8, 12.4] | 11.9 | [10.4, 13.5] | 8.4 | [7.1, 9.6] | 8.0 | [7.1, 8.9] | 7.0 | [6.2, 7.9] |
| Left home | 28.2 | [20.1, 36.4] | 64.8 | [62.6, 67.1] | 79.5 | [77.6, 81.3] | 70.3 | [68.7, 71.8] | 67.2 | [65.7, 68.7] |
| Avoid crowds | 88.8 | [83.1, 94.5] | 51.3 | [48.9, 53.6] | 70.6 | [68.6, 72.7] | 64.8 | [63.2, 66.4] | 74.8 | [73.4, 76.2] |
| Avoid transport | 42.2 | [33.3, 51.2] | 32.0 | [29.8, 34.2] | 48.6 | [46.3, 50.8] | 50.0 | [48.3, 51.6] | 49.0 | [47.4, 50.6] |
| Use facemask | 20.7 | [13.3, 28.1] | 73.5 | [71.4, 75.6] | 95.7 | [94.7, 96.6] | 94.5 | [93.7, 95.2] | 97.6 | [97.1, 98.1] |
| Unable access medication | 45.8 | [36.8, 54.8] | 45.7 | [43.4, 48.1] | 48.2 | [45.9, 50.4] | 58.0 | [56.4, 59.7] | 54.3 | [52.7, 55.9] |
| Unmet healthcare need | 11.0 | [5.4, 16.7] | 10.5 | [9.0, 11.9] | 5.6 | [4.6, 6.7] | 6.6 | [5.8, 7.4] | 5.5 | [4.7, 6.2] |
| Lost earnings | 50.0 | [41.0, 59.0] | 28.8 | [26.7, 31.0] | 16.3 | [14.6, 18.0] | 15.6 | [14.4, 16.8] | 13.3 | [12.2, 14.4] |
| PHQ-2 | 16.9 | [10.2, 23.7] | 7.8 | [6.5, 9.1] | 3.0 | [2.2, 3.7] | 12.3 | [11.2, 13.4] | 8.8 | [7.9, 9.8] |
| GAD-2 | 28.0 | [19.9, 36.1] | 8.5 | [7.2, 9.8] | 6.3 | [5.2, 7.4] | 13.3 | [12.2, 14.4] | 11.0 | [10.0, 12.0] |

## Notes

### Competing Interest Statement

The authors have declared no competing interest.

### Author Declarations

All study procedures were discussed with each nodes' existing community advisory groups and approved by Limpopo, Mpumalanga and KwaZulu-Natal's provincial Department of Health Research Ethics Committees (REC), the University of KwaZulu-Natal's Biomedical REC, the University of the Witwatersrand's Human REC (Medical) and the University of Limpopo's Turfloop REC.

